# The impact of natural disasters on the spread of COVID-19: a geospatial, agent based epidemiology model

**DOI:** 10.1101/2020.09.12.20193433

**Authors:** Maximillian Van Wyk de Vries, Lekaashree Rambabu

## Abstract

**Background:** Natural disasters and infectious diseases are global issues, resulting in widespread disruption to human health and livelihood. At the scale of a global pandemic, the cooccurrence of natural disasters is inevitable. However, the impact of natural disasters on the spread of COVID-19 has not been extensively evaluated using agent based epidemiology models.

**Methods:** We create an agent-based epidemiology model based on both COVID-19 clinical and epidemiological data and geographic data. We first model 35 scenarios with varying natural disaster timing and duration for a COVID-19 outbreak in a theoretical region. We then evaluate the potential effect of an eruption of Vesuvius volcano on the spread of COVID-19 in Campania, Italy. Our objective is to determine if the occurrence of a natural disaster during the COVID-19 pandemic is likely to increase infection cases and disease related fatalities.

**Results:** In a majority of cases, the occurrence of a natural disaster increases the number of disease related fatalities. When the natural disaster occurs at the beginning of the outbreak within a given region, there is little to no increase in the progression of disease spread. However, the occurrence of a natural disaster close to the peak of infections may increase the number of fatalities by more than five-fold. In a theoretical test case, for a natural disaster that occurred fifty days after first infection case, the median increase in fatalities is 2%, 59%, and 180% for a 2, 14, and 31-day long natural disaster respectively, when compared to the no natural disaster scenario.

**Conclusion:** We propose that the compound risk from natural disasters is greatest in the case of already widespread disease outbreak. The key risk factors for increase in spread of infection and disease related fatalities are the timing of the natural disaster relative to the peak in infections and the duration of the natural disaster.

## Introduction

As of September 2020, COVID-19 has spread to over 200 countries, infected more than 28.9 million, and killed over 920,000 people [1]. Natural disasters can threaten the measures in place to reduce disease transmission [2]. Understanding how disease spread can be worsened by natural disasters will aid preparedness and response planning. Although a small increase in risk of epidemic outbreak following natural disasters has been identified [2, 3], very little is known about the spread of infection following a natural disaster during a pandemic such as COVID-19.

Numerical modelling is an important tool for understanding and projecting the spread of infectious diseases [4], including COVID-19 [5, 6, 7]. A common approach is to divide a population up into susceptible, infected, recovered and deceased individuals [4, 8]. More sophisticated models incorporate a larger number of different states (e.g. asymptomatic or seriously ill individuals; [5]), and may account for other real world complications (e.g. age, vaccination campaigns, etc.).

The time varying proportion of each state may be calculated deterministically by solving a series of ordinary differential equations [4, 5]. This approach can accurately portray the spread of infectious diseases on a large scale [4, 5]. The deterministic approach is ill suited for simulation of small population sizes, heterogeneous populations and threshold behaviour [7]. A stochastic, agent based model structure may also be used [8, 9], in which case the variation between runs allows for quantification of uncertainty [6]. Agent based models are are well suited to represent heterogeneous or complex populations and spatial variability [8, 9].

## Methods

We use geoSIR, a geospatial, agent based model structure to account for the large scale movement of individuals within the model space from a natural disaster evacuation [8, 9]. Each cell may be in one of 7 states: susceptible (*S*), infected and mildly ill (*I_m_*), infected and severely ill (*I_s_*), infected and asymptomatic (*I_a_*), recovered (*R*), deceased (*D*) or empty (*X_e_*). Empty space allows us to account for other geographic factors such as population density. Individuals may also self isolate or be quarantined. We base the definition of our states and model disease characteristics on COVID-19 data [10, 11, 12, 13, 14, 15].

In the initial state, all individuals are susceptible. The spread of the disease is initiated with the arrival of infected travellers. Two parameters may be adjusted to control the seeding of the disease: the number of incoming travellers (*Tr_in_*) and the probability that each traveller is infected (*P_TrI_*). The number of imported cases per day *C_I_* is:

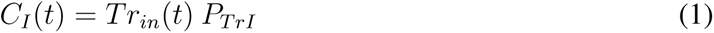

For the majority of the world, new COVID-19 cases were initially imported by a small number of travellers before spreading throughout the community [16]. As individuals are infected, the number of susceptible individuals 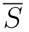 evolves as:

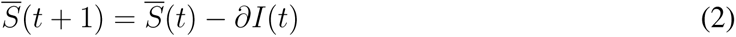

With *∂I* the number of new infected individuals in day *t*. The total number of new infected individuals depends on the total number of infected individuals in the previous day 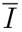, and a disease spread parameter *α*:

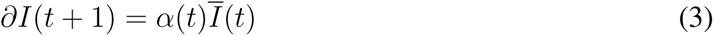

With *α* given by:

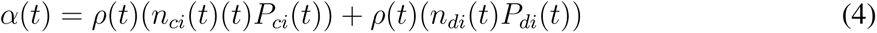

With *n_ci_* the number of close encounters, *P_ci_* the probability of infection for close encounters, *n_di_* the number of distant encounters, and *P_di_* the probability of infection for close encounters. Close encounters can represent sustained contact with another individual within a household or among close friends. Distant encounters can represent an individual’s risk of catching an infection through indirect contact such as visits to supermarkets, fomites transmission or whilst commuting. The details of these two parameters can be modified to account for specific modes of transmission. We use data from two case studies of COVID-19 transmission in Wenzhou, China [17] and Bavaria, Germany [15] to estimate parameters. We also ensure that resulting reproductive number *R*_0_ (as calculated from equation 6) are consistent with the range estimated for COVID-19 [10, 14]. We use 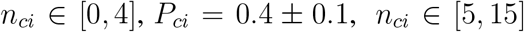, and 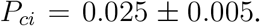. The number of close and distant encounters are comparable to those used for an agent based simulation of influenza outbreaks in New York City [8].

The number of encounters and probability of infection transmission can be reduced through lockdowns and social distancing measures. Equally, they can be increased by large gatherings or movements of people. *ρ* is a measure of the local proportion of individuals that are susceptible:

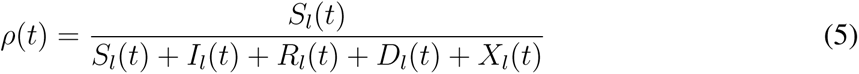

*S_I_, I_I_, R_I_, D_I_*,and *X_I_* being respectively the number of susceptible, infected, recovered, dead and empty spaces in the relevant local travel region *l*. The radius *l* is larger for distant encounters than for close encounters in order to account for spread outside of an individual’s direct social circle. The mean reproductive number *R*_0_(*t*) of the disease at time *t* is calculated as:

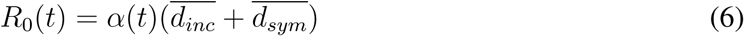

With 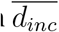 the mean incubation time and 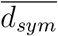 the mean duration of symptoms. We use 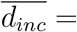 days, and 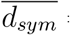 days [12, 18, 13]. A bulk *R*_0_ of the disease is calculated at each time step, however internal variability within the simulated region can be high. This can result in periods of time in which the regional average reproductive number is less than 1, however the infection still spreads rapidly in local hotspots. Estimates of reproductive number are influenced by the local mitigation practices, environment and individual behaviour and comparison between regions is complex [19].

The daily change in total number of infected individuals is:

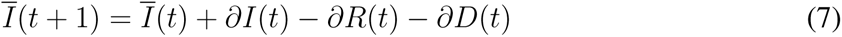

Where *∂R* is the new number of recovered individuals in a given day, and *∂D* the number of new deaths. 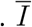 includes asymptomatic, mild and severe cases in varying stages of the disease.

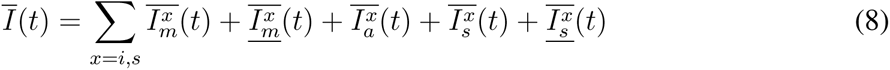

A proportion *P_A_* of infections begin as asymptomatic, and the remaining 1 *- P_A_* begin as mild. The proportion of asymptomatic COVID-19 patients has been estimated at around 20% through airport screenings [20]. We use *P_A_* = 0.2 *±* 0.05. Asymptomatic infected individuals may infect others, but do not change state until they recover. Mild cases have a *P_S_* chance of transitioning to severe cases over the course of their illness (*d_sym_*). We use a definition of severe cases that combines the ‘severe’ and ‘critical’ categories from Wu and McGoowan (2020), resulting in *P_S_* = 0.19 *±* 0.02 [11]. Each severely ill individual has a probability *P_D_* of dying. The expected probability of dying for any infected individual, termed infection fatality rate *P_IFR_* is thus:

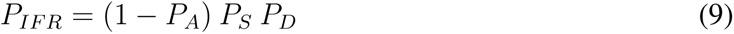

Many of the regions worst affected by COVID-19 experienced their highest death rates at times when hospital capacity was reached or exceeded [21]. A hospital capacity term *H_C_* allows for the simulation of scenarios in which hospital capacity becomes overwhelmed. This may represent number of intensive care unit (ICU) beds, number of ventilators or some other measure of how many severely ill patients can be treated at any given time. We also include a term that describes the probability of a seriously ill individual being hospitalised *P_H_*. The death rate is for seriously ill individuals is given by:

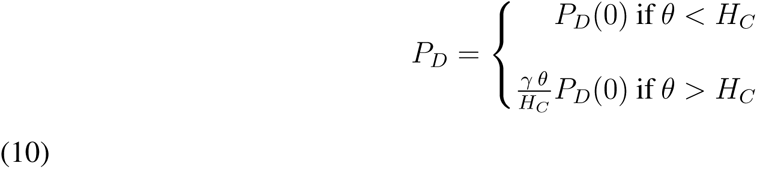

With *P_D_*(0) the base death rate for seriously ill patients, *γ* a scaling factor for increases in death rate and

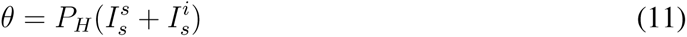

A death rate of severely ill patients is capped at *P_D_*(*max*).

We use COVID-19 infection mortality rates from from a compilation of Chinese data [11,12], calculated to be 2.3% and 1.4% respectively. We define *P_D_*(0) = 0.12 *±* 0.02, giving an approximate infection mortality rate of 1.8%. We define hospital capacity according to local ICU bed capacities.

If an infected person does not die after the duration of their symptoms (*d_sym_*), they recover. This model does not account for possible re-infection, and recovered individuals remain immune for the duration of the model run. The simulation of testing is possible in this model (see user manual), but we make the simplifying assumption that testing has not had a major impact on COVID-19 mitigation in the regions considered.

The model simulation grid may be built according to real-world geospatial data. We vary the initial proportion of susceptible individuals *S* and empty space *X_e_* to account for differences in population density. High and medium natural hazard areas are also inputted. These determine the areas in which evacuation may be necessary. For Vesuvius, high and medium hazard zone masks are created based on the ‘red’ and ‘yellow’ zones defined in the most recent Vesuvius National Emergency Plan [22]. Individuals living in high or medium hazard zones are tagged, and retain more social interactions (e.g. a higher number of close or distant encounters) than other individuals following evacuation. Social distancing is challenging both during evacuation [23].

Information on changes in mobility following natural disasters is limited. Lu et al. (2012) analysed mobile phone data trends following the 2010 Haiti earthquake. They found that the number of people travelling more than 20 km increased 86% in the day following the earthquake, and that increase in movement lasted two to three weeks[24]. These movements led to a 23% decrease in the population of capital city Port-au-Prince, equivalent to close to half a million outward journeys [24]. Lockdowns will be temporarily interrupted during evacuations, and many individuals will be unable to maintain social distancing or lockdown rules [2]. To account for this, we increase the number of encounters by a factor of 3 for individuals within the high and medium risk zones, and by a factor of 1.5 in the low risk zone. We model two different scenarios: an idealised region containing a small city and concentric geological hazard, and a theoretical eruption of Vesuvius during the COVID-19 pandemic in Campania, Italy.

### Theoretical region

In order to quantify disease spread during natural disasters in general, we create a non-specific experimental region. This region includes an idealised town or city with high population densities that radially decay away from the town centre. Initial effective population densities 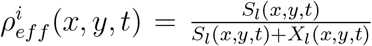 vary from 1 for the centre of town to 0.3 in rural areas. A concentric geological hazard is located within this study area, with the medium hazard region overlapping moderate population density outskirts of the city and rural areas. Hazard *N_H_* is divided into high and medium hazard zones, with the high hazard zone entirely enveloped by the medium hazard zone. The theoretical region has a population of 100,000 individuals, and a total number of cells equal to 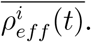. We find that a population of 100,000 is sufficiently large to accurately reproduce disease spread, yet low enough to remain computationally efficient. We conduct 100 different runs for each scenario, seeding the Mercenne Twister with system time to ensure independent runs.

### Campania

Campania, Italy has a population of 5.8 m inhabitants, includes Italy’s third largest city (Napoli), and to active volcano Vesuvius. With eight Plinian eruptions in the last 25,000 years and 32 confirmed eruption in the last 1000 years [25], further eruptions in the near future are likely. The potential damage associated with a large eruption of Vesuvius is severe. However, comprehensive risk mitigation strategies have been derived for the volcanic hazards [26]. The key mitigation strategy is a timely evacuation [26]. We investigate the effect of the evacuation of a large area around Vesuvius during an infectious disease outbreak similar to COVID-19, with the assumption that no one is directly harmed by the volcanic eruption. This disaster response results in widespread population displacement - a key risk factor in disease spread [2].

We use published data for the spread of COVID-19 to parametrise our model, and compare the outputs of the most realistic scenario (no eruption, lockdown initiated) to real world daily infection and fatality data. We use the number of ICU beds in Campania region (427) as a measure for hospital capacity [27]. Due to the large population size in this scenario (over 5 million individuals), we adapt the code to run on a 24 core Haswell E5-2680v3 processor node of the Minnesota Supercomputing Institute.

## Results

In the theoretical region, we perform 100 model runs for 35 different scenarios. These scenarios cover various permutations of lockdown, natural disaster timing and natural disaster duration. Both the number of infections and number of deaths are higher when a natural disaster occurs during the infection outbreak (Fig. 1). The scale of this increase depends on the relative timing of the natural disaster and peak in infection cases, and on the duration of the disaster (Figure 1). In the case with no natural disaster, the median number of infections is 4,900 (IQR 4,140-5,640) cases per 100,000 and the median number of deaths is 66 (IQR 53-76) per 100,000. The increase in deaths remains low when the natural disaster occurs during the onset of the outbreak (day 1, median 57, IQR 39-100 deaths per 100,000) and after the initial infection peak has subsided (day 200, median 73, IQR 53-92.5 deaths per 100,000 respectively). The number of infections and deaths is highest when the natural disaster occurs close to the peak of the outbreak. In this case, the number of infections are 153% higher and deaths are 602% higher when compared to the scenario with no natural disaster (NDt = day 20, median 12,390, IQR 7,100-16,820 cases per 100,000; median 463, IQR 126-847 deaths per 100,000).

**Figure 1:**
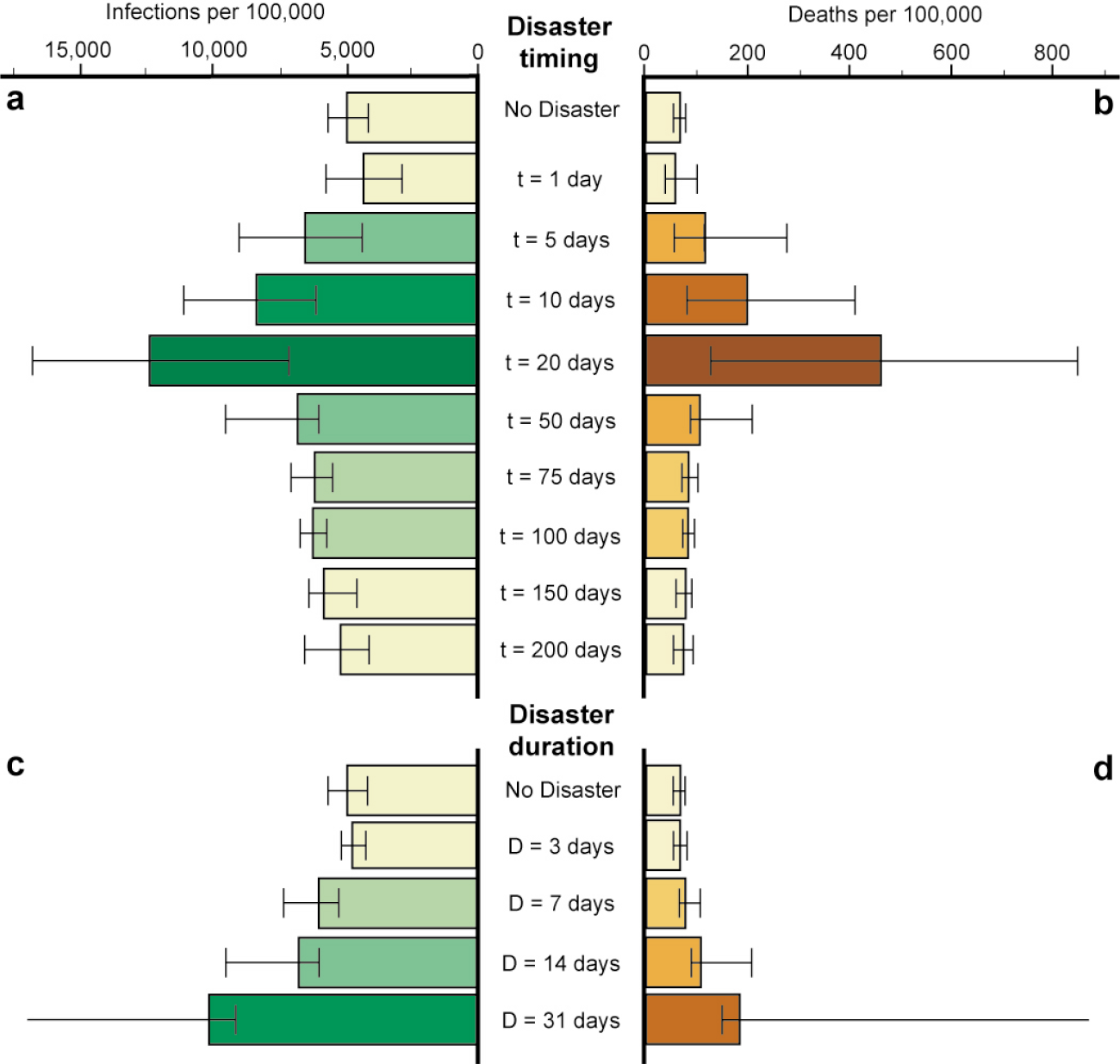
Number of infections and deaths for different theoretical disasters. The timing (**a** and **b**) and duration (**c** and **d**) of the ND are varied. Natural disasters occuring close to the peak of infections (around 20–30 days) have the largest impact. The increase in infections and deaths increases non-linearly with the duration of the disaster.

For a given natural disaster timing (50 days), long disruptions cause more infections and deaths than short ones. A 3 day long natural disaster results in a similar number of deaths (median 66.5, IQR 55-80.5 per 100,000) as the case with no natural disaster. A 31 day long natural disaster, however, causes and 180% increase in median number of deaths (median 183.5 IQR 149.5-3172 deaths per 100,000). In the case where no lockdown is instigated, around 80% of the population is infected and death toll is extreme (median 3028, IQR 2674-3259 deaths per 100,000).

For Campania, we first model the COVID-19 outbreak in a scenario similar to reality (lock-down initiated, no Vesuvius eruption). Our model predicts a median of 580 (IQR 553-641) infections per 100,000, and a median of 8.5 (IQR 7.9-8.9) deaths per 100,000. Model infections are lower, but consistent with Vollmer et al., (2020)’s estimate of 590-950 cases per 100,000 [28] and model deaths are consistent with real world data (7.5 deaths per 100,000 as of July 2020) [1].

We then model five counterfactual scenarios, four of which include an eruption of Vesuvius (Figure 2) during Campania’s COVID-19 outbreak. The increase in total number of deaths remains low for an early (Day 2, mean 9.6, IQR 7.6-13.1 deaths per 100,000) or post infections peak (Day 100, median 10.9, IQR 10-12.1 deaths per 100,000) Vesuvius eruption. The median deaths rise to 68.5 (IQR 12.1-133.3) per 100,000 when a Vesuvius eruption occurs close to the infection peak (Day 50). In this case, the deaths per 100,000 are comparable to the states in Northern Italy worst hit by COVID-19 (168, 95 and 44 deaths per 100,000 in Lombardy, Piedmont and Veneto respectively as of September 2020) or other hard-hit regions such at New York State, USA (168 deaths per 100,000 as of September 2020) [1]. For an eruption occurring close to the peak in infections at day 50, there is a large variability in possible outcomes. The median death rate is 8 times higher than with no eruption (68.5 deaths per 100,000). However, in 32% of runs the death rate is no more than double that of the no eruption scenario, while in 12% of runs the death rate is more than 25 times higher.

**Figure 2:**
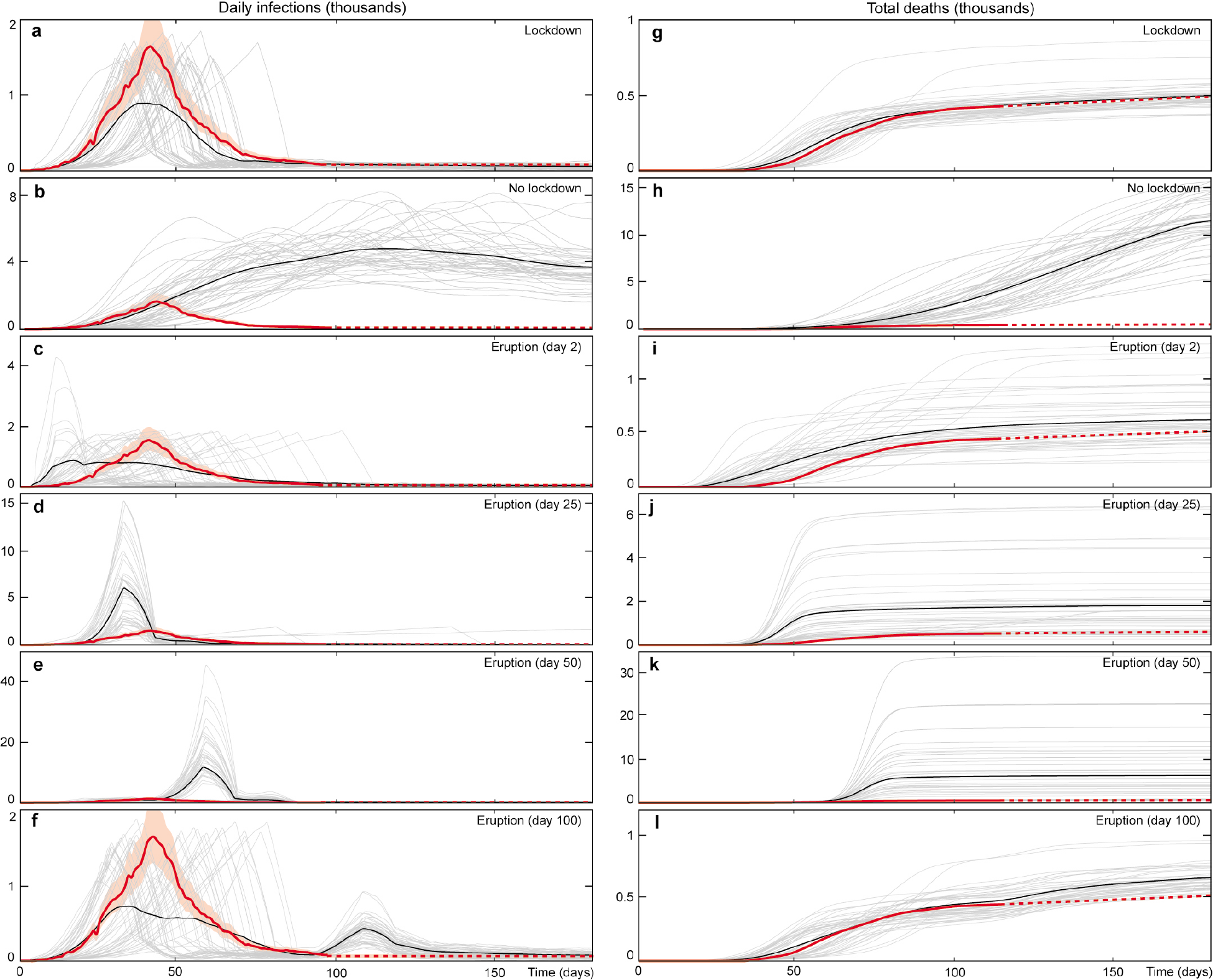
Results of the number of daily infections and deaths from COVID-19 in Campania, Italy under different scenarios. The red line represents real world data, the black line represents model mean and the grey lines represent individual model runs. All values are filtered with a 10 day moving mean to remove short period noise. The in the scenario with no eruption and a lockdown, model outputs are close to the observed real world data (**a** and **g**). Note that the model mean new daily cases appear artificially low due to different peak timings, but that the magnitude of individual runs are comparable to the real world outputs. The mean number of deaths is higher in all scenarios in which Vesuvius erupts (**c** and **i, d** and **j, e** and **k, f** and **l**,), and higher by more than an order of magnitude where the eruption coincides with the peak in infections (**e** and **k**).

Italy was one of the most hard hit countries during the COVID-19 pandemic [5], with over 33,000 deaths and 233,000 confirmed cases as of June 2020. Campania had more than 4800 confirmed infections and 400 deaths [1]. Although the probability of Vesuvius erupting during the COVID-19 pandemic is low, the wealth of volcanic evacuation plans and COVID-19 data make it a valuable case study. The exact nature of the geological hazard is not of primary importance, rather the parameters of the evacuation and increased human contact following influence the progression of the infectious disease outbreak. The Campania results may be used to understand the effect of other disasters requiring widespread evacuation during a pandemic.

Natural disasters have already occurred during the COVID-19 pandemic [29, 30], and will occur during future infectious disease outbreaks. Cyclone Amphan made landfall in Bangladesh and West Bengal, India on May 20, 2020, prompting widespread evacuations. The number of new COVID-19 infections in the first week of June was 3.5 times higher in West Bengal and 4.8 times higher in Bangladesh compared to the same period in May [1]. The number of COVID-19 related deaths rose by a similar percentage. Further research is required to determine whether Cyclone Amphan played a role in this increase in infections and deaths, which at this stage cannot be attributed to any particular cause.

The majority of previous studies on the relationship between infectious disease outbreak and natural disasters have focused on whether natural disasters initiate new outbreaks. In a small number of cases, a disease outbreak has followed a natural disaster [2]. However, a review of the topic concluded that “the risk for epidemics after a geophysical disaster is very low” [3]. Our results do not contradict this, but highlight the previously overlooked extreme risk in cases of already widespread infection.

**Figure 3:**
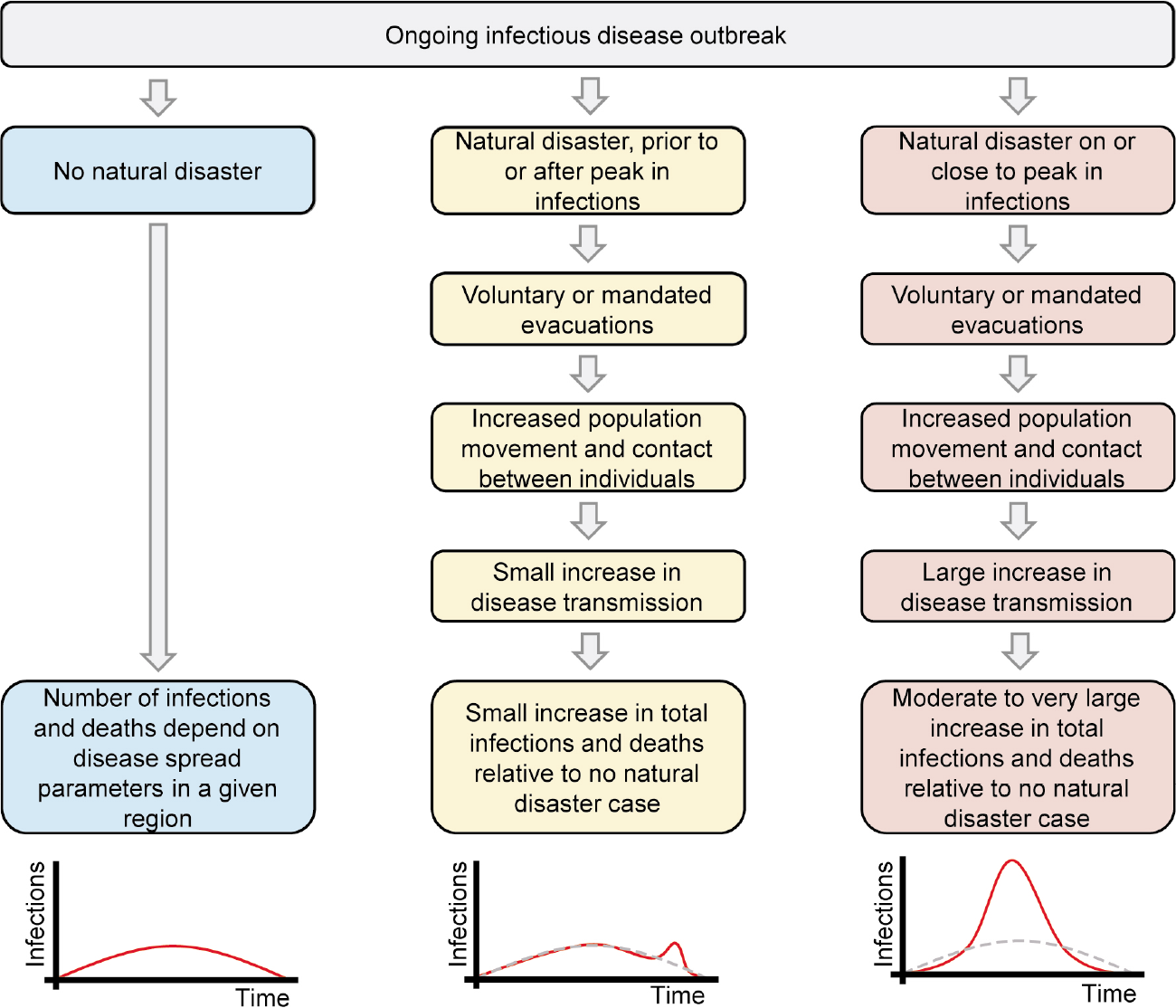
Summary of impact of natural disasters on disease outbreak, as modelled in this study.

The possibility of natural hazards interacting with COVID-19 has previously been raised [29, 30]. Quigley et al., 2020 use a phenomenological model to investigate the impact of natural disaster occurrence during the COVID-19 pandemic. They also find that this results in a larger total number of infections, although the simplicity of their model precludes detailed interpretation [29]. Phillips et al., 2020 propose that climate change is increasing the magnitude and frequency of climatic hazards, therefore raising the risk of natural disasters co-occurring with disease outbreaks [30]. Overall, our results show that the co-occurrence of a natural disaster increases COVID-19 spread. Furthermore, we demonstrate that the timing and duration of the natural disaster are key risk factors for increase in infections. Close links between the epidemiology response and natural disaster response communities are necessary for the formulation of timely risk assessments.

## Data Availability

All relevant data is available upon contact with the corresponding author.

## References

[1] Dong, E., Du, H. & Gardner, L. An interactive web-based dashboard to track COVID-19 in real time. The Lancet Infectious Diseases 20, 533–534 (2020). URL http://www.sciencedirect.com/science/article/pii/S1473309920301201.

[2] Kouadio, I. K., Aljunid, S., Kamigaki, T., Hammad, K. & Oshitani, H. Infectious diseases following natural disasters: prevention and control measures. Expert Review of Anti-Infective Therapy 10, 95–104 (2012).

[3] Floret, N., Viel, J.-F., Mauny, F., Hoen, B. & Piarroux, R. Negligible Risk for Epidemics after Geophysical Disasters. Emerging Infectious Diseases 12, 543–548 (2006). URL https://www.ncbi.nlm.nih.gov/pmc/articles/PMC3294713/.

[4] Brauer, F. Compartmental Models in Epidemiology. Mathematical Epidemiology 1945, 19–79 (2008). URL https://www.ncbi.nlm.nih.gov/pmc/articles/PMC7122373/.

[5] Giordano, G. et al. Modelling the COVID-19 epidemic and implementation of population-wide interventions in Italy. Nature Medicine 1–6 (2020). URL http://www.nature.com/articles/s41591-020-0883-7. Publisher: Nature Publishing Group.

[6] Hellewell, J. et al. Feasibility of controlling COVID-19 outbreaks by isolation of cases and contacts. The Lancet Global Health 8, e488–e496 (2020). Publisher: Elsevier.

[7] Kucharski, A.J. et al. Early dynamics of transmission and control of COVID-19: a mathematical modelling study. The Lancet Infectious Diseases 20, 553–558 (2020). URL http://www.sciencedirect.com/science/article/pii/S1473309920301444.

[8] Cooley, P. et al. The Role of Subway Travel in an Influenza Epidemic: A New York City Simulation. Journal of Urban Health: Bul-letin of the New York Academy of Medicine 88, 982–995 (2011). URL https://www.ncbi.nlm.nih.gov/pmc/articles/PMC3191213/.

[9] Hackl, J. & Dubernet, T. Epidemic Spreading in Urban Areas Using Agent-Based Transportation Models. Future Internet 11, 92 (2019). URL https://www.mdpi.com/1999-5903/11/4/92. Number: 4 Publisher: Multidisciplinary Digital Publishing Institute.

[10] Flaxman, S. et al. Estimating the effects of non-pharmaceutical interventions on COVID-19 in Europe. Nature 1–8 (2020). URL http://www.nature.com/articles/s41586-020-2405-7. Publisher: Nature Publishing Group.

[11] Wu, Z. & McGoogan, J.M. Characteristics of and Important Lessons From the Coronavirus Disease 2019 (COVID-19) Outbreak in China: Summary of a Report of 72 314 Cases From the Chinese Center for Disease Control and Prevention. JAMA 323, 1239–1242 (2020). URL https://jamanetwork.com/journals/jama/fullarticle/2762130. Publisher: American Medical Association.

[12] Guan, W.-j. et al. Clinical Characteristics of Coronavirus Disease 2019 in China. New England Journal of Medicine (2020). URL https://www.nejm.org/doi/10.1056/NEJMoa2002032. Publisher: Massachusetts Medical Society.

[13] Zhou, F. et al. Clinical course and risk factors for mortality of adult inpatients with COVID-19 in Wuhan, China: a retrospective cohort study. The Lancet 395, 1054–1062 (2020). URL https://www.thelancet.com/journals/lancet/article/PIIS0140-6736(20)30566-3/abstract Publisher: Elsevier.

[14] Liu, Y., Gayle, A.A., Wilder-Smith, A. & Rocklöv, J. The reproductive number of COVID-19 is higher compared to SARS coronavirus. Journal of Travel Medicine 27 (2020). URL https://academic.oup.com/jtm/article/27/2/taaa021/5735319. Publisher: Oxford Academic.

[15] Böhmer, M. M. et al. Investigation of a COVID-19 outbreak in Germany resulting from a single travel-associated primary case: a case series. The Lancet Infectious Diseases 0 (2020). URL https://www.thelancet.com/journals/laninf/article/PIIS1473-3099(20)30314-5/abstract Publisher: Elsevier.

[16] Chinazzi, M. et al. The effect of travel restrictions on the spread of the 2019 novel coronavirus (COVID-19) outbreak. Science 368, 395–400 (2020). URL https://science.sciencemag.org/content/368/6489/395. Publisher: American Association for the Advancement of Science Section: Research Article.

[17] Cai, J. et al. Indirect Virus Transmission in Cluster of COVID-19 Cases, Wenzhou, China, 2020 – Volume 26, Number 6—June 2020 - Emerging Infectious Diseases journal – CDC (2020). URL https://www.nc.cdc.gov/eid/article/26/6/20-0412article.

[18] Lauer, S.A. et al. The Incubation Period of Coronavirus Disease 2019 (COVID-19) From Publicly Reported Confirmed Cases: Estimation and Application. Annals of Internal Medicine 172, 577–582 (2020). URL https://www.acpjournals.org/doi/full/10.7326/M20-0504. Publisher: American College of Physicians.

[19] Delamater, P.L., Street, E.J., Leslie, T.F., Yang, Y.T. & Jacobsen, K. H. Complexity of the Basic Reproduction Number (R0). Emerging Infectious Diseases 25 (2020). URL https://www.nc.cdc.gov/eid/article/25/1/17-1901article.

[20] Quilty, B.J., Clifford, S., Group2, C.n. w., Flasche, S. & Eggo, R.M. Effectiveness of airport screening at detecting travellers infected with novel coronavirus (2019-nCoV). Eurosurveillance 25, 2000080 (2020). URL https://www.eurosurveillance.org/content/10.2807/1560-7917.ES.2020.25.5.2000080 Publisher: European Centre for Disease Prevention and Control.

[21] Boccia, S., Ricciardi, W. & Ioannidis, J. P.A. What Other Countries Can Learn From Italy During the COVID-19 Pandemic. JAMA Internal Medicine (2020). URL https://jamanetwork.com/journals/jamainternalmedicine/fullarticle/2764369

[22] De Vivo, B. & Rolandi, G. Vesuvius: volcanic hazard and civil defense. Rendiconti Lincei 24, 39–45 (2013). URL https://doi.org/10.1007/s12210-012-0212-2.

[23] Tanigawa, K., Hosoi, Y., Hirohashi, N., Iwasaki, Y. & Kamiya, K. Loss of life after evacuation: lessons learned from the Fukushima accident. The Lancet 379, 889–891 (2012). URL https://www.thelancet.com/journals/lancet/article/PIIS0140-6736(12)60384-5/abstract Publisher: Elsevier.

[24] Lu, X., Bengtsson, L. & Holme, P. Predictability of population displacement after the 2010 Haiti earthquake. Proceedings of the National Academy of Sciences 109, 11576–11581 (2012). URL http://www.pnas.org/cgi/doi/10.1073/pnas.1203882109.

[25] Rolandi, G., Paone, A., Di Lascio, M. & Stefani, G. The 79 AD eruption of Somma: The relationship between the date of the eruption and the southeast tephra dispersion. Journal of Volcanology and Geothermal Research 169, 87–98 (2008). URL http://www.sciencedirect.com/science/article/pii/S037702730700279X.

[26] Baxter, P.J. et al. Emergency planning and mitigation at Vesuvius: A new evidence-based approach. Journal of Volcanology and Geothermal Research 178, 454–473 (2008). URL http://www.sciencedirect.com/science/article/pii/S0377027308004630.

[27] Pecoraro, F., Clemente, F. & Luizi, D. The efficiency in the ordinary hospital bed management in Italy: an in-depth analysis of intensive care unit in the areas affected by COVID-19 before the outbreak | medRxiv. medRxiv preprint (2020). URL https://www.medrxiv.org/content/10.1101/2020.04.06.20055848v1.

[28] Vollmer, M. et al. Report 20: A sub-national analysis of the rate of transmission of Covid-19 in Italy. Tech. Rep., Imperial College London (2020). URL http://spiral.imperial.ac.uk/handle/10044/1/78677.

[29] Quigley, M.C., Attanayake, J., King, A. & Prideaux, F. A multi-hazards earth science perspective on the COVID-19 pandemic: the potential for concurrent and cascading crises. Environment Systems & Decisions 1–17 (2020). URL https://www.ncbi.nlm.nih.gov/pmc/articles/PMC7229439/.

[30] Phillips, C. A. et al. Compound climate risks in the COVID-19 pandemic. Nature Climate Change 1–3 (2020). URL http://www.nature.com/articles/s41558-020-0804-2. Publisher: Nature Publishing Group.

